# Novel Investigation of SARS-CoV-2 in COVID-19 Survivors’ Semen in Surabaya, Indonesia

**DOI:** 10.1101/2021.10.08.21264593

**Authors:** Supardi Supardi, Reviany V. Nidom, Eni M. Sisca, Jefry A. Tribowo, Patricia S. Kandar, Joice M. Budiharto, Eko Siswidiyanto, Maitra Djiang Wen, Tiara Kirana, Astria N. Nidom, Arif N. M. Ansori, Irine Normalina, Setyarina Indrasari, Reny I’tishom

## Abstract

The emergence and the widespread of Coronavirus disease 2019 (COVID-19) demands an accurate detection method to establish a diagnosis. Real-time polymerase chain reaction (real-time PCR) is accounted for the perfect point of reference in detecting this virus. The notion that this virus also invades the male reproductive tract requires further investigation to prove the presence of severe acute respiratory syndrome coronavirus-2 (SARS-CoV-2) in the semen. This investigation was designed to detect SARS-CoV-2 in COVID-19 survivors’ semen. This study design was a cross-sectional examination and conducted between November 2020 and March 2021 in the Andrology Unit of Dr. Soetomo General Hospital and Professor Nidom Foundation, both located in the City of Surabaya, Indonesia. The sample was 34 male participants aged above 18 years old and had been confirmed COVID-19 by nasopharyngeal swab PCR test. Part of the semen was taken for real-time PCR testing with the QuantStudio 5 Applied Biosystem (AB) PCR machine and the kits utilized were the STANDARD M nCOV Real-Time Detection Kit and mBioCov-19 RT-PCR Kit. Furthermore, the mean of participants’ ages was 35.74 years old with 25% of them had had a history of primary infertility and 21.8% of secondary infertility. From the real-time PCR COVID-19 of the semen examination, this investigation found that 27 participants had been negatives (74.4%), six inconclusive (17.6%), and one positive (3%) of SARS CoV-2. In summary, SARS-CoV-2 could be found in the semen of COVID-19 survivors. This should be a concern for the potential impact of COVID-19 in male fertility and the possibility of transmission reproductively.

## 1. Introduction

COVID-19 occurs as the result of contagiousness from severe acute respiratory syndrome coronavirus-2 (SARS-CoV-2) spread abruptly and became a global pandemic since December 2019 as the first report of the emergence of this virus in Wuhan [1,2]. As of June 2021, there is around 170 million confirmed prevalence of COVID 19 infection globally with more than 3.5 million global deaths. To be specific, Indonesia has about 1.8 million cases and around 50 thousand deaths [1,2,3,4,5]. The involvement of this disease causes a very broad impact and clinical manifestations that vary, ranging from asymptomatic to systemic dysfunction [6,7]. Furthermore, SARS-CoV-2 involves angiotensin II converting enzymes (ACE2) as a receptor for entering into the cells. This expression of ACE 2 is found in the intestine, kidney, and testicle [8]. Although ACE2 expression is present in the testes, the direct involvement of SARS-CoV-2 infection in the testes or male genital tract is still in question [9].

Various testing approaches were developed to detect this virus. An accurate and fast detection method is needed as part of preventive and curative efforts [10]. Until recently, three types of diagnostic tests have been developed, namely reverse transcription-polymerase chain reaction (RT-PCR) assay, antibody detection and tissue culture isolation [11]. The most highly recommended, which puts a highly accurate benchmark, uses a nucleic acid-based approach such as the polymerase chain reaction (PCR) method as it has rapid, sensitive, and specific virus detection capabilities, even PCR can detect early SARS-CoV-2 infection. PCR analysis utilizes specimens generated from saliva; the swabbing activity of nasal, trachea and nasopharyngeal; tissues of lungs; blood; semen; and even; excrement. [12,13]. The extraction process of ribonucleic acid (RNA) in the specimen starts from the purification process of total RNA into viral RNA and host RNA, then continued by reversed transcription into the complementary DNA (cDNA) and followed by amplification using the polymerization of chain reaction which is the foundation of real time reverse transcription-polymerase chain reaction (rRT-PCR) examination. [14]. Although rRT-PCR is the most widely used and recommended as the barometer of SARS COV-2 diagnostics, it also has drawbacks. Errors in the preparation, analysis, and interpretation stages will give false positive or false negative or inconclusive rRT-PCR results [15,16].

Unlike rRT-PCR examination of nasopharyngeal specimens which is routinely performed, the use of rRT-PCR in semen specimens is still limited and is more for research purposes. There were various studies discussing COVID-19 rRT-PCR in semen [11,13,15,16,17]. Among these various studies, only two studies detected SARS-CoV-2 in the semen [18,19]. Therefore, this investigation was the novel finding in Indonesia to examine the semen of male COVID-19 survivors using rRT-PCR test method to confirm whether SARS-COV-2 was found in the men reproductive tract of the survivors.

## 2. Materials and Methods

### 2.1. Design study and participants

This research is a cross-sectional examination. This investigation was conducted between November 2020 and March 2021 in the Andrology Unit of Dr. Soetomo Regional Hospital and in the Professor Nidom Foundation, both located in Surabaya, Indonesia. A total of 34 male participants were registered as research subjects. Inclusion criteria were males older than 18 years old, had been confirmed COVID-19 by nasopharyngeal PCR test, and already in the recovery phase. This research was ethically evaluated with an ethical exemption with the reference number of 274/EC/KEPK/FKUA/2020 from the Faculty of Medicine, Universitas Airlangga.

### 2.2. Data collection

This investigation performed historical consideration and semen analysis on the participants who met the inclusion criteria. Participants were then instructed to abstinence 2-7 days and wash their hands before doing masturbation for the semen collection.

### 2.3. Real-Time PCR

The examination was implemented using RT-PCR with QuantStudio 5 Applied Biosystem (AB) PCR machine, the kit applied was STANDARD M nCOV Real-Time Detection Kit (Lot #MNCO0120026) and MBioCoV-19 RT-PCR Kit (Lot # 6900820) carried out at the Research Group of Coronavirus and Vaccine Formulation, at Professor Nidom Foundation, Surabaya, Indonesia. This investigation utilized a kit based on Taqman probe real time fluorescent PCR technology. The foundation of PCR examination was the transcription of coronavirus RNA into cDNA by reverse transcriptase as the first step, then cDNA was employed as a framework for PCR amplification.

PCR reaction simultaneously applied activities of polymerase such as DNA polymerase and exo-nuclease. The detachment of both fluorophore and quencher along with the activity of dicer, which has the capability to discredit the TaqMan probe, provoke the instrument to indicate the signal of fluorescence; for example, new coronavirus ORP1ab (RdRp) gene was detected by FAM channel qualitative, coronavirus E gene was identified by JOE (VIC or HEX) channel qualitative indication, and internal reference was recognized by CY5 channel detection. In preventing the contamination of amplification products, dUTP and UNG enzymes were employed by this kit. Positive criteria were detected if 2 gene (E and RdRP) ≤36. If the results only found in E gene ≤36, it meant that the inconclusive. Negative criteria are E and RdRP not detected.

Moreover, PCR examination was carried out in two stages. In the first stage, PCR examination of semen sample code 1-15 was performed on January, 2021 and in the second stage, PCR for semen sample code 16-34 was implemented on March, 2021. Before the rRT PCR examination was conducted, the specimens were stored in a refrigerator at 4°C.

### 2.4. Statistical analysis

The data from anamnesis and the interpretation of PCR results that had been collected were analyzed and grouped by demographics, the rRT-PCR results, the rRT-PCR results based on the degree of symptoms, the duration of time between nasopharyngeal PCR and semen PCR, and lastly, the symptoms.

## 3. Results

In this investigation, the rate of male COVID-19 survivors’ age was 37.60±7.834, while for their spouses was 37.44±5.525. The average duration of marriage was 9.11 years with 90% of them were married. Furthermore, the primary infertility rate was 44.4%, and, for secondary infertility, it was 22.2% (Table 1). In the results of real-time PCR COVID-19 in the semen, it was found that 27 participants were negative (74.4%), 6 participants were inconclusive (17.6%), and 1 participant was positive (3%) (Table 2 and Figure 1).

**Table 1:** Demographic Characteristics of Participants

| Variable |  |
| --- | --- |
| Total participants (n) | 10 |
| Mean age | $37.60 \pm 7.834$ |
| Mean couples age | $37.44 \pm 5.525$ |
| Mean marriage duration | 9.11 |
| Marital Status |  |
| Married | 9 (90%) |
| Not Married | 1 (10%) |
| Primary Infertility |  |
| Yes | 4 (44.4%) |
| No | 5 (55.6%) |
| Secondary Infertility |  |
| Yes | 2 (22.2%) |
| No | 7 (77.8%) |

**Table 2:** Semen rRT-PCR Results.

| Code | E | RdRP | IC | Interpretation |
| --- | --- | --- | --- | --- |
| S1 | - | - | 23.91 | Negative |
| S2 | - | - | 21.11 | Negative |
| S3 | - | - | 26.07 | Negative |
| S4 | - | - | 21.24 | Negative |
| S5 | - | - | 20.21 | Negative |
| S6 | - | - | 20.49 | Negative |
| S7 | - | - | 21.95 | Negative |
| S8 | - | - | 25.59 | Negative |
| S9 | - | - | 22.87 | Negative |
| S10 | 32.64 | 36.16 | 23.31 | Positive |
| S11 | - | - | 26.58 | Negative |
| S12 | - | - | 27.96 | Negative |
| S13 | - | - | 25.29 | Negative |
| S14 | - | - | 24,23 | Negative |
| S15 | - | - | 26.41 | Negative |
| S16 | - | - | 26.88 | Negative |
| S17 | - | - | 19.35 | Negative |
| S18 | - | - | 21.11 | Negative |
| S19 | - | 35.86 | 21.57 | Inconclusive |
| S20 | - | - | 22.9 | Negative |
| S21 | - | 35.99 | 20.47 | Inconclusive |
| S22 | 33.77 | 42.62 | 19.36 | Inconclusive |
| S23 | - | 35.64 | 23.78 | Inconclusive |
| S24 | - | 34.99 | 17.52 | Inconclusive |
| S25 | - | 27.65 | 21.19 | Inconclusive |
| S26 | - | - | 24.51 | Negative |
| S27 | - | - | 24.05 | Negative |
| S28 | - | - | 26.45 | Negative |
| S29 | - | - | 27.47 | Negative |
| S30 | - | - | 23.5 | Negative |
| S31 | - | - | 25.33 | Negative |
| S32 | - | - | 24.43 | Negative |
| S33 | - | - | 24.91 | Negative |
| S34 | - | - | 26.86 | Negative |

**Figure 1.**
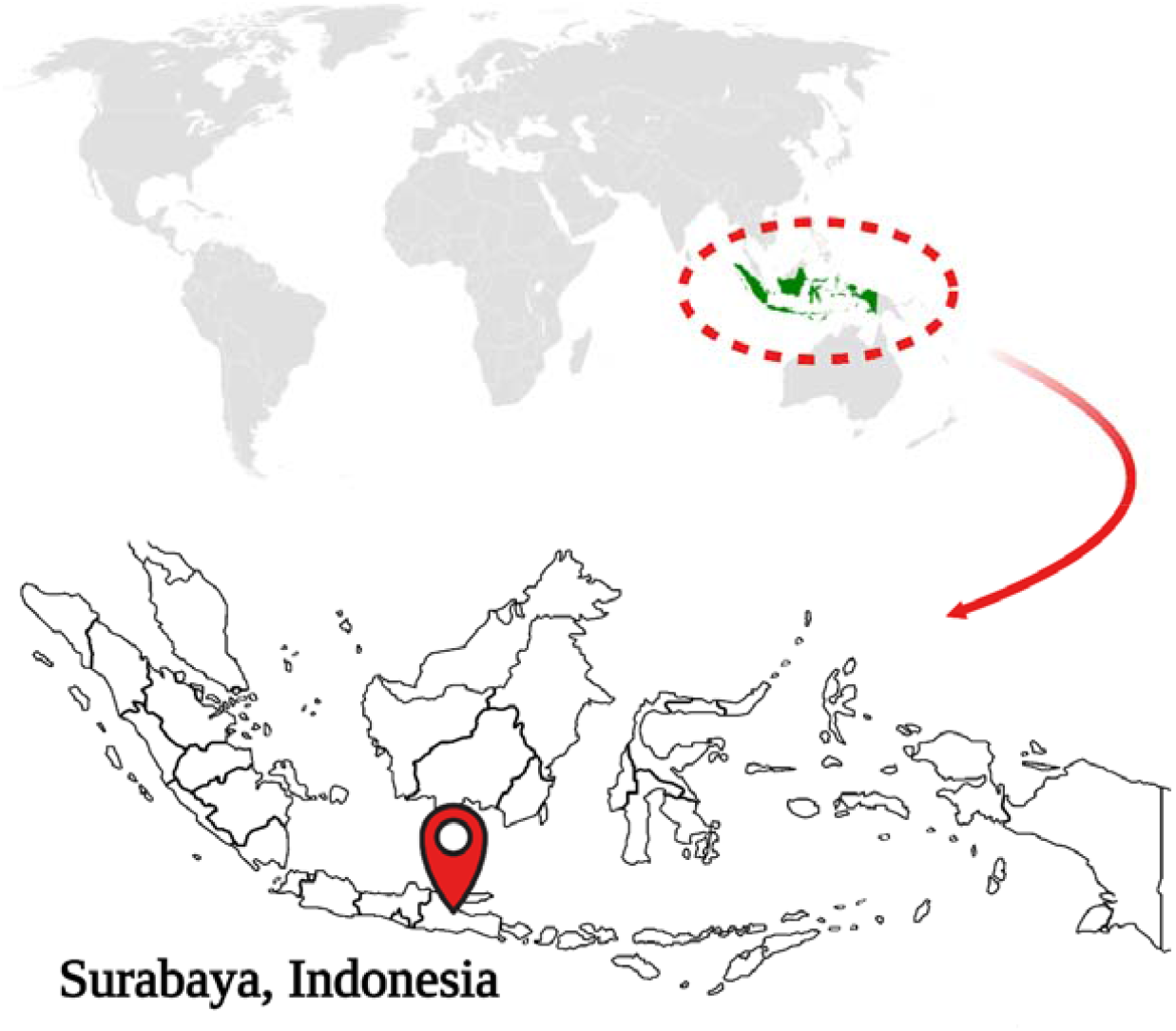
Location of the Study. This study conducted in the Andrology Unit, Dr.Soetomo Regional Hospital Surabaya and Professor Nidom Foundation, Surabaya, Indonesia.

Meanwhile, rRT-PCR examinations based on the degree of COVID-19 symptoms were classified into 4 groups, they were asymptomatic with negative semen rRT-PCR result in 13 participants (38.23%), asymptomatic COVID-19 with positive semen rRT-PCR result in 1 participant (2.94%), mild symptoms with negative semen rRT-PCR result in 11 participants (32.35%), mild symptoms with inconclusive semen rRT-PCR result in 6 participants (17.64%), and moderate symptoms with negative semen rRT-PCR result in 3 participants (8.82%) (Table 3 and Figure 2).

**Table 3:** Interval Duration of rRT-PCR Examination in Nasopharyngeal and Semen

| Sample | rRT PCR Semen (n = 7) | rRT PCR Nasopharyngeal | Interval Duration |
| --- | --- | --- | --- |
| S10 | Positive | Positive | 121 days |
| S19 | Inconclusive | Positive | 177 days |
| S21 | Inconclusive | Positive | 126 days |
| S22 | Inconclusive | Positive | 59 days |
| S23 | Inconclusive | Positive | 186 days |
| S24 | Inconclusive | Positive | 53 days |
| S25 | Inconclusive | Positive | 71 days |
| Total |  |  | 793 days |
| Mean |  |  | 113.28 days |

**Figure 2.**
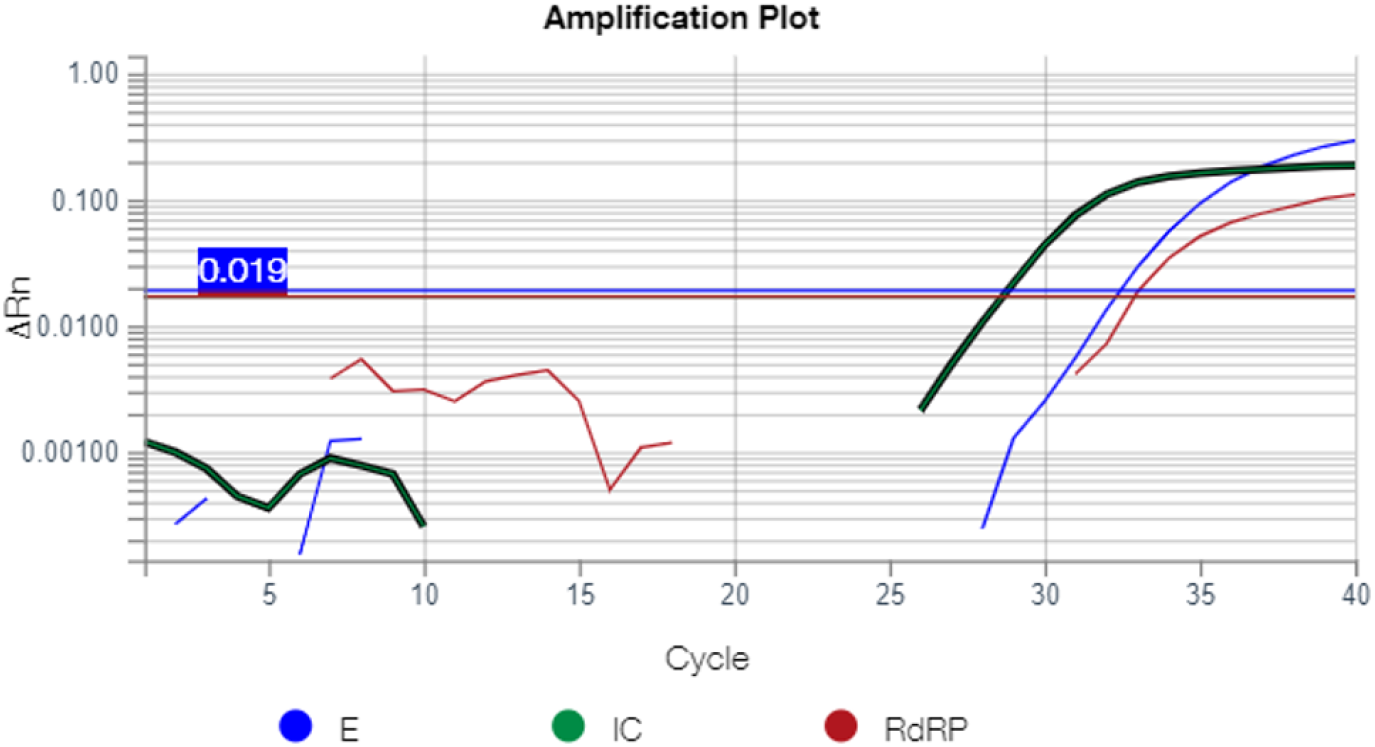
rRT-PCR of S10 with IC: 23.31.

Furthermore, the average of interval duration between positive or inconclusive rRT for nasopharyngeal PCR and rRT for semen PCR with positive or inconclusive results was 113.28 days. The fastest interval duration between positive and inconclusive nasopharyngeal PCR and semen PCR was 53 days, while the longest interval duration to produce positive or inconclusive PCR was 186 days (Table 4).

**Table 4.**
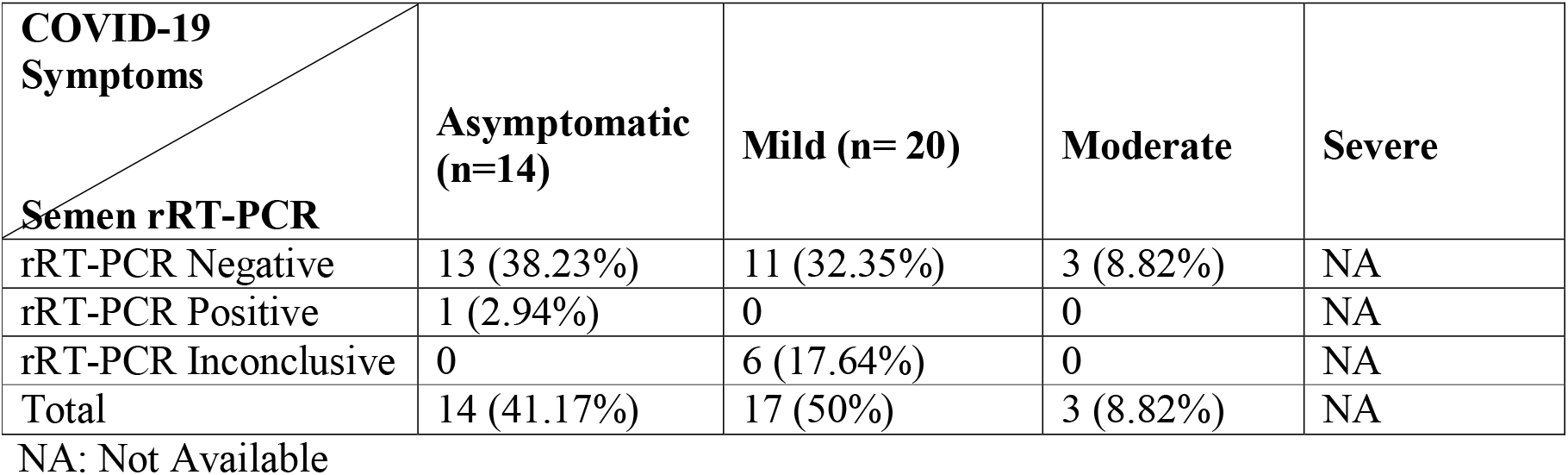
Distribution of PCR results based on disease severity.

Based on the severity classification of COVID-19 according to WHO [18], this study distributed the symptoms of COVID 19 into asymptomatic, mild, moderate and severe COVID 19 (Table 5). There were not any participants in this study who had a severe symptom of COVID-19.

**Table 5:** COVID-19 symptoms of participants

| Symptoms | Total (n=34) |
| --- | --- |
| Asymptomatic | 14 (41.18%) |
| Mild |  |
| Anosmia | 8 (23.53%) |
| Hypogeusia | 1 (2.94%) |
| Flu-like Syndrome (fever, cough, cold, and sore throat) | 10 (29.41%) |
| Gastrointestinal Symptoms (vomit and diarrhea) | 2 (5.88%) |
| Insomnia | 1 (2.94%) |
| Orchitis | 1 (2.94%) |
| Moderate |  |
| Shortness of Breath | 3 (8.82%) |
| Pneumonia on Chest X-ray | 3 (8.82%) |

## 4. Discussion

In this examination, as the mean of participants’ age was 37.7 years, the oldest was about 50’s years old and the youngest was about 20’s years old. Most of the participators were sexually active and had children. Age is one of the prognostic factors of participants infected with COVID-19, as the older is the age, the greater is the possible risk of worsening symptoms [20,21,22].

Meanwhile, the results of the semen PCR in this examination were inconclusive for 6 participants (17.6%). In Gacci *et al*. (2021) research, they revealed one participant was inconclusive [18], while the study of Bhattacharya *et al*. (2020) stated that the results of rRT nasopharyngeal swab were inconclusive around 5%. There are several factors that affect the inconclusive PCR results from the virologic perspective; due to RNA extraction errors, inadequate sample quality (internal housekeeping genes were not detected in RT-PCR), other beta corona virus infections, very early or late stage of infection, differences in analytic sensitivity in E and S gene or RdRP, distinction in the specificity of screening and confirming gene PCRs. In addition, it can be caused as well because of the improper preanalytical stage in the form of sampling and no cold chain provision leading to RNA degradation, poor quality of transporter medium, poor storage space, and lack of amplification in internal control or housekeeping genes [23].

Correspondingly, there was only one participant with positive semen rRT-PCR in this study. The time span between positive nasopharyngeal rRT-PCR and positive semen rRT-PCR in this participant was 121 days. This participant was about 30’s years old, married, and had 2 children. He had asymptomatic on COVID-19 and no history of any systematic disease. He had central obesity according to the Asian-Pacific criteria [24].

Furthermore, Li et al (2020) stated that SARS COV-2 was found in semen in two positive participants based rRT PCR examination in the recovery phase, while other 4 participants were detected positive in the semen during the acute phase. In this study, the discovery of one participant who was positive for SARS COV-2 in the semen has similarities with the study of Gacci *et al*. (2021) where there was only one participant who was positive for SARS COV-2 in the semen, but when the nasopharyngeal PCR test was evaluated, the results were negative [18]. Whereas in this investigation, a nasopharyngeal PCR for reevaluation was not performed on this patient.

Additionally, one participant in this study complained on the testicular pain at initial symptom onset. In other study, it was found there were 6 cases of orchitis from the autopsy of SARS patients [25]. This spread in the reproductive tract is probably related to viral loads in the body and related to the infectious process (Figure 3 and Figure 4) [26].

**Figure 3:**
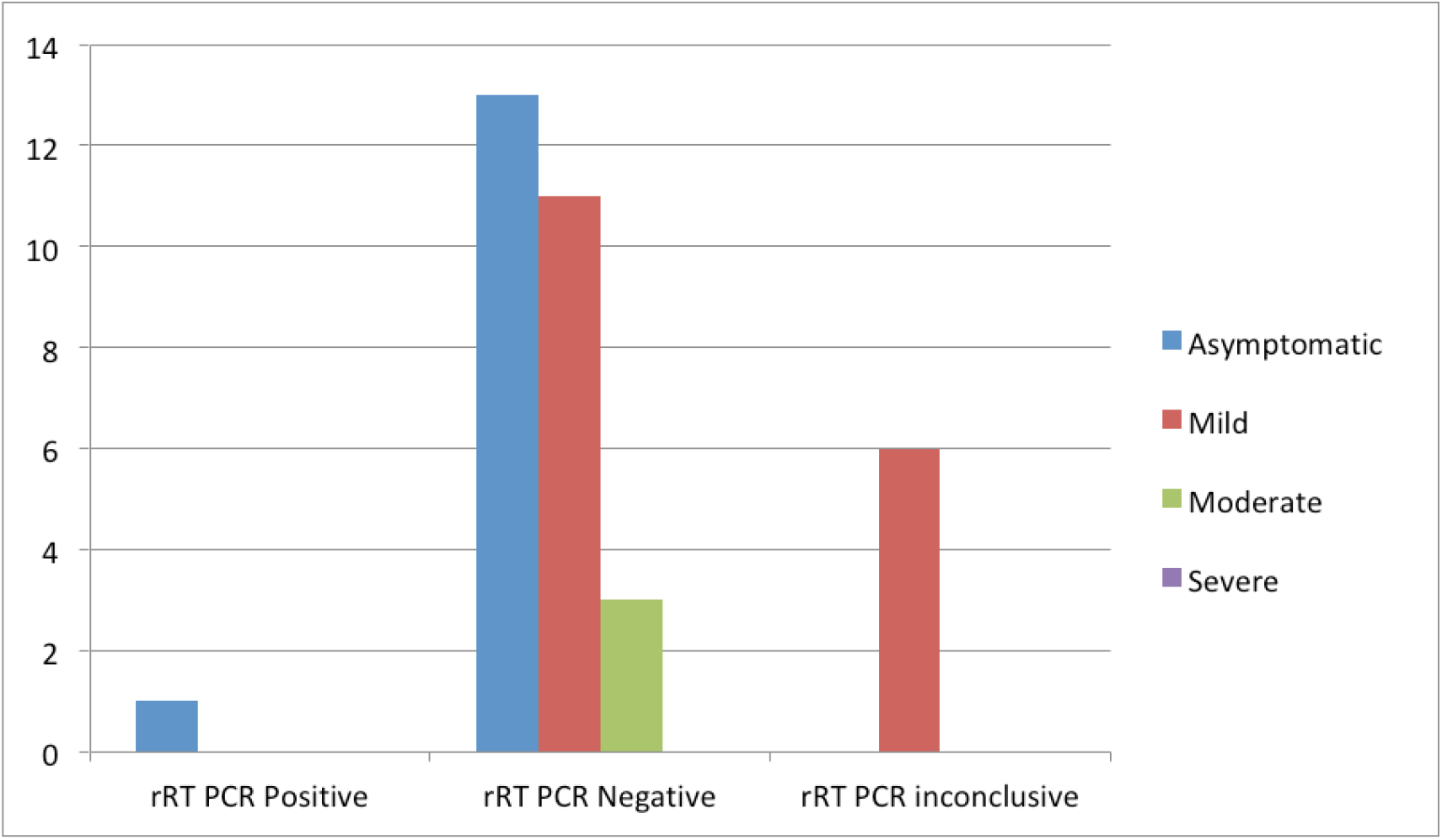
Distribution of rRT-PCR results based on disease severity

**Figure 4:**
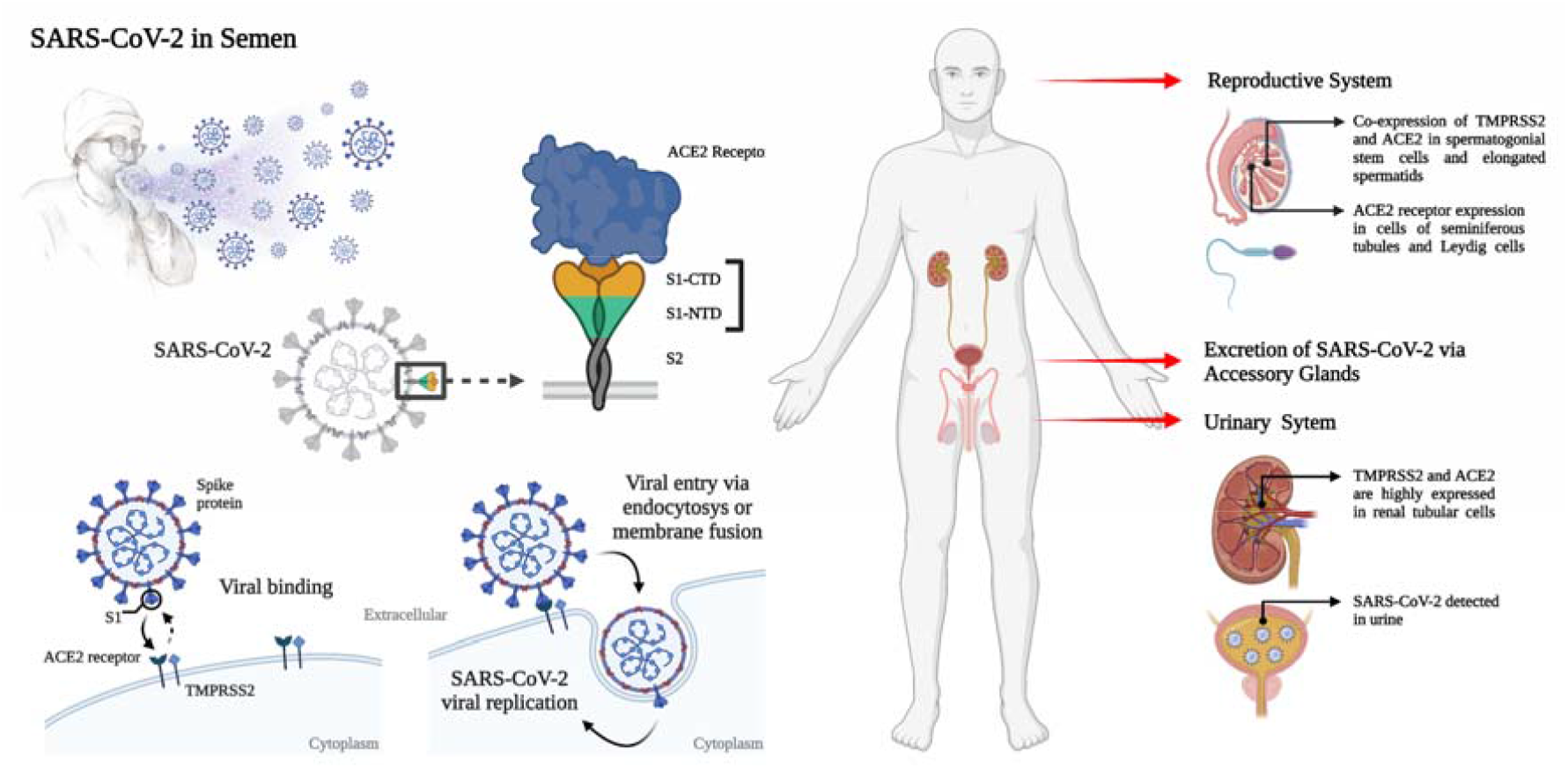
Feasible scenarios and viral entry action for SARS-CoV-2 in semen. Infection of this virus is dependent on both TMPRSS2 and ACE2 expression for the entry into the host cell. This figure created with BioRender.

Since most of the participants were detected negative for semen PCR (74.4%), this was probably because the recruited participants had already passed the recovery phase. Furthermore, the results of rRT PCR were more often negative because there was only <1% chance of detecting positivity in the semen [15,16].

Finally, the weakness in this current investigation was that the transport of the samples did not use a cold chain box, thus, there was an opportunity for errors occurring at the pre-analytic stage. This PCR examination was not examined when the sample first received, instead it was stored in the refrigerator for several days. Participants with positive PCR results in semen were no longer confirmed by nasopharyngeal PCR, therefore, it was not known whether there was a recurrent positive.

## 5. Conclusions

In summary, SARS-CoV-2 could be found in the semen of COVID-19 survivors. It should be noted that PCR is influenced by preanalytical, analytical, and interpretive circumstances. The SARS-CoV-2 existence in the semen sample requires to be further evaluated in future study, as this could be a concern for the consequences of COVID-19 on male fertility status and the possibility of sexual transmission.

## Data Availability

The utilized data to contribute to this investigation are available from the corresponding author on reasonable request.

## Data Availability

The utilized data to contribute in this investigation are available from the corresponding author on reasonable request.

## Acknowledgments

The authors would like to convey gratitude to all employees at the Andrology Unit, Dr. Soetomo Regional Hospital Surabaya and the Department of Biomedical Science in the Faculty of Medicine of Universitas Airlangga, specifically to all participators, colleagues, and acquaintances contributed in this study for their tremendous assistance to complete this investigation. This research was fully supported by Universitas Airlangga’s grant for COVID-19 Research (Grant Number: 664/UN.3.14/PT/2020) and the Professor Nidom Foundation, Surabaya, Indonesia (Grant Number: 006/PNF-RF/10/2020).

